# Optimising primary molecular profiling in NSCLC

**DOI:** 10.1101/2023.08.20.23294346

**Authors:** R.D. Schouten, I. Schouten, M.M.F. Schuurbiers, V. van der Noort, R.A.M. Damhuis, E.H.F.M. van der Heijden, J.A. Burgers, N.P. Barlo, A.S.R. van Lindert, K.W. Maas, J.J.G. van den Brand, A.A.J. Smit, J.M.W. van Haarst, B. van der Maat, E. Schuuring, H. Blaauwgeers, S.M. Willems, K. Monkhorst, D. van den Broek, M.M. van den Heuvel

## Abstract

**Introduction:** Molecular profiling of NSCLC is essential for optimising treatment decisions, but often incomplete. We assessed the efficacy of protocolised molecular profiling in the current standard-of-care (SoC) in a prospective observational study in the Netherlands and measured the effect of providing standardised diagnostic procedures. We also explored the potential of plasma-based molecular profiling in the primary diagnostic setting.

**Methods:** This multi-centre prospective study was designed to explore the performance of current clinical practice during the run-in phase using local SoC tissue profiling procedures. The subsequent phase was designed to investigate the extent to which comprehensive molecular profiling (CMP) can be maximized by protocolising tumour profiling. Successful molecular profiling was defined as completion of at least *EGFR* and *ALK* testing. Additionally, PD-L1 tumour proportions scores were explored. Lastly, the additional value of centralised plasma-based testing for *EGFR* and *KRAS* mutations using droplet digital PCR was evaluated.

**Results:** Total accrual was 878 patients, 22.0% had squamous cell carcinoma and 78.0% had non-squamous NSCLC. Stage I-III was seen in 54.0%, stage IV in 46.0%. Profiling of *EGFR* and *ALK* was performed in 69.9% of 136 patients included in the run-in phase, significantly more than real-world data estimates of 55% (*p*<0.001). Protocolised molecular profiling increased the rate to 77.0% (*p*=0.049). *EGFR* and *ALK* profiling rates increased from 77.9% to 82.1% in non-squamous NSCLC and from 43.8% to 57.5% in squamous NSCLC. Plasma-based testing was feasible in 98.4% and identified oncogenic driver mutations in 7.1% of patients for whom tissue profiling was unfeasible.

**Conclusion:** This study shows a high success rate of tissue-based molecular profiling that was significantly improved by a protocolised approach. Tissue-based profiling remains unfeasible for a substantial proportion of patients. Combined analysis of tumour tissue and circulating tumour DNA is a promising approach to allow adequate molecular profiling of more patients.

## Introduction

Biomarker testing in patients with metastatic non-small cell lung cancer (NSCLC) is essential for making optimal treatment decisions.(1–3) There is an increasing number of targetable alterations for which tyrosine kinase inhibitors (TKIs) are available(4), which demands comprehensive molecular profiling (CMP). When there is no targeted treatment eligible, PD-L1 testing guides the choice of treatment. Moreover, recent studies have shown that various biomarkers are becoming relevant in early stages of NSCLC as well(5–7), which underlines the importance of CMP for all patients with NSCLC.

However, molecular analysis of lung cancer is hampered by various problems. Tumour biopsies are invasive and not without risk of complications. The yield of tissue biopsies is typically small, while the requirements for tissue increase with the number of tests needed to assess the variety of molecular alterations.(8–10) Additionally, multiple testing requires multiple iterations of tissue preparation. This often results in loss of tissue, further reducing the availability of valuable tumour material. Besides the issue of unavailable or unsuitable material for CMP, current diagnostic workup regularly does not include all relevant molecular targets. Previous studies have shown an increased rate of molecular testing in recent years, but further improvement is still warranted.(11,12)

The development of circulating tumour DNA (ctDNA) analysis has shown great progress in recent years. These blood-based tests have the potential to overcome some of the issues encountered with tissue-based tests. The minimally-invasive nature makes it suitable for almost any patient and may be of additional value to current molecular profiling. However, clinical validity in NSCLC has yet to be determined.(13,14)

We primarily aimed to explore how much molecular diagnostics would improve by protocolising molecular testing. At the time of study design only testing for *EGFR* and *ALK* in stage IV NSCLC were considered standard of care (SoC) in the Netherlands. Additionally, this study design proposed CMP should include other biomarkers as well (*KRAS, BRAF, ERBB2, ROS1, RET, MET*, and PD-L1). Moreover, we envisioned molecular profiling of early stage disease would become clinically relevant. Therefore, this study is an exploration of biomarker prevalence in all histological types and all stages of NSCLC. Furthermore, this study explores the feasibility and added value of ctDNA analysis by droplet digital PCR (ddPCR) in the primary diagnostic setting.

## Methods

### Study design, setting and subjects

In this multicentre prospective cohort study executed in ten hospitals across The Netherlands (Supplementary Table 1) patients with a suspicion of non-small cell lung cancer (NSCLC) or established NSCLC but still awaiting treatment were recruited. Data were collected longitudinally during the recruitment period that started on August 1^st^ 2016 and ended on December 31^st^ 2019. All patients provided written informed consent to participate in this diagnostic study. The only exclusion criterion was if the patient was not willing to undergo any form of treatment. Patients were considered screen failures when no malignancy was found, when they were diagnosed with another malignancy than NSCLC, when they were not treatment-naïve before collection of tumour tissue and blood, and when they had already been diagnosed with NSCLC in the five years leading up to this study. The LEMA study protocol is available at ClinicalTrials.gov (NCT02894853).

### Study procedures

The run-in phase was designed to explore the performance of current routine clinical practice and simultaneously explore the effect of increased awareness on molecular profiling. In this phase, participating centres were allowed to perform molecular profiling according to the local SoC. Duration was maximised to six months and ended for all centres before January of 2019.

In the protocol phase, CMP was protocolised for all patients with NSCLC, independent of stage and histology. Tumour tissue had to be processed to allow all biomarker analyses at once, thereby minimising loss of tissue. Testing for oncogenic driver variants in *EGFR, KRAS, BRAF, ERBB2, ALK, ROS1, RET* and *MET*, and assessment of PD-L1 expression were obligatory. All tissue-based biomarker assessments were performed locally in the centres of enrolment. The panel of biomarkers was selected based on contemporary national and international guidelines and recommendations. Patients received treatment according to the SoC or were included in clinical studies where appropriate.

During both study phases, blood samples from all patients were collected before the start of treatment. Blood samples were centrally stored and processed. Samples from patients at the site of the central laboratory were collected in standard K2-EDTA-tubes. Samples from other participating sites were collected in cell-stabilizing tubes (STRECK, Omaha, USA) and sent to the central laboratory. Blood samples were centrifuged for 10 minutes at 1700G at room temperature. Cells were stored at -80°C and plasma was centrifuged for 10 minutes at 20,000G before storage at -80°C. Analysis of ctDNA with ddPCR was performed on 4 ml of plasma per patient and included *EGFR* exon 19 deletions, *EGFR* L858R and T790M mutations, and KRAS mutations (G12A/C/D/R/S/V and G13D). For isolation of cell-free DNA (cfDNA) the QIAsymphony Circulating DNA kit (article number 1091063, Qiagen, Düsseldorf, Germany) with QIAsymphony (Qiagen) was used.

### Statistical considerations

The LEMA study is a unique project in the Netherlands. It is the first large, multicentre study designed to improve molecular diagnostics in NSCLC. During the design of the study, various parameters that were necessary to calculate sample- and effect sizes were unknown. Therefore, several assumptions had to be made and it was expected that these parameters needed adjustment after data from the LEMA cohort itself became available. A detailed description of the sample- and effect size calculation is provided in the supplement.

To quantify the effect of the LEMA protocol we postulated that the rate of testing for *EGFR* and *ALK* should exceed 55% to be clinically relevant. Statistical significance was assessed by calculating the Z-statistic. Statistical significance of the increased testing rate in the protocolised phase compared to the run-in phase was assessed by Fisher’s exact test. Since testing and identification rates were expected to increase only, a one-sided alpha of <0.05 was considered to be the threshold for significance.

### Ethical statement

The Lung Cancer Early Molecular Assessment trial (LEMA) was reviewed and approved by the medical ethics committee of the Netherlands Cancer Institute in Amsterdam, The Netherlands (Clinicaltrials.gov identifier NCT 02894853). The study was conducted in accordance with the Declaration of Helsinki (as revised in 2013) and the guidelines for Good Clinical Practice.

## Results

### Cohort characteristics

A total of 1108 patients were assessed for eligibility in ten participating hospitals in the Netherlands and 230 patients were excluded. During the run-in phase of the study 136 patients were included. The protocolised phase of the LEMA trial included 742 patients. The duration of inclusion was 40 months, between September 2016 and December 2019. Cohort characteristics, reasons for ineligibility and distribution across study phases are shown in table 1 and figure 1.

**Figure 1.**
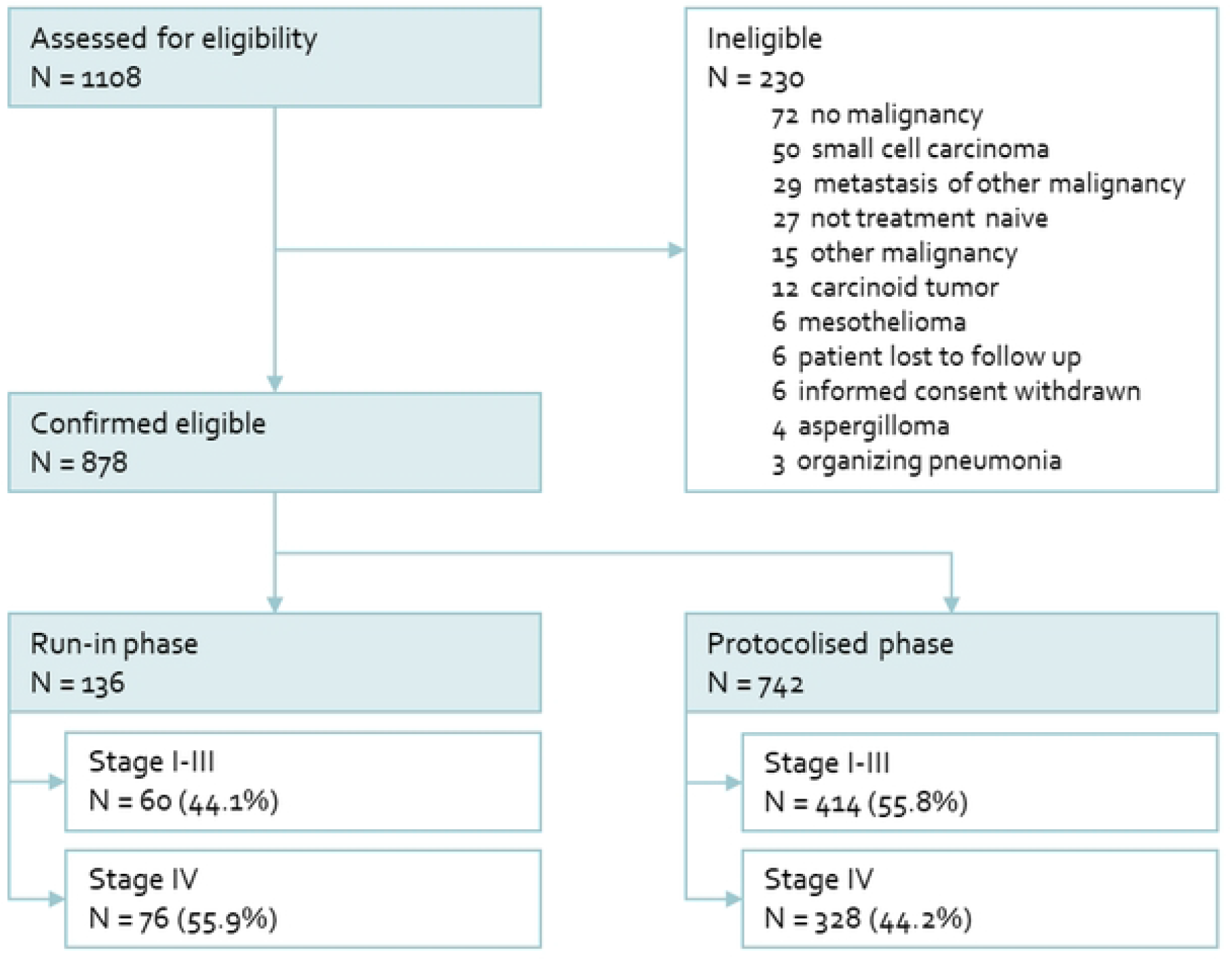
Inclusion flow chart. Reasons for ineligibility are shown in the top right panel. A more detailed flow chart is shown in supplementary figure S1.

**Table 1.**
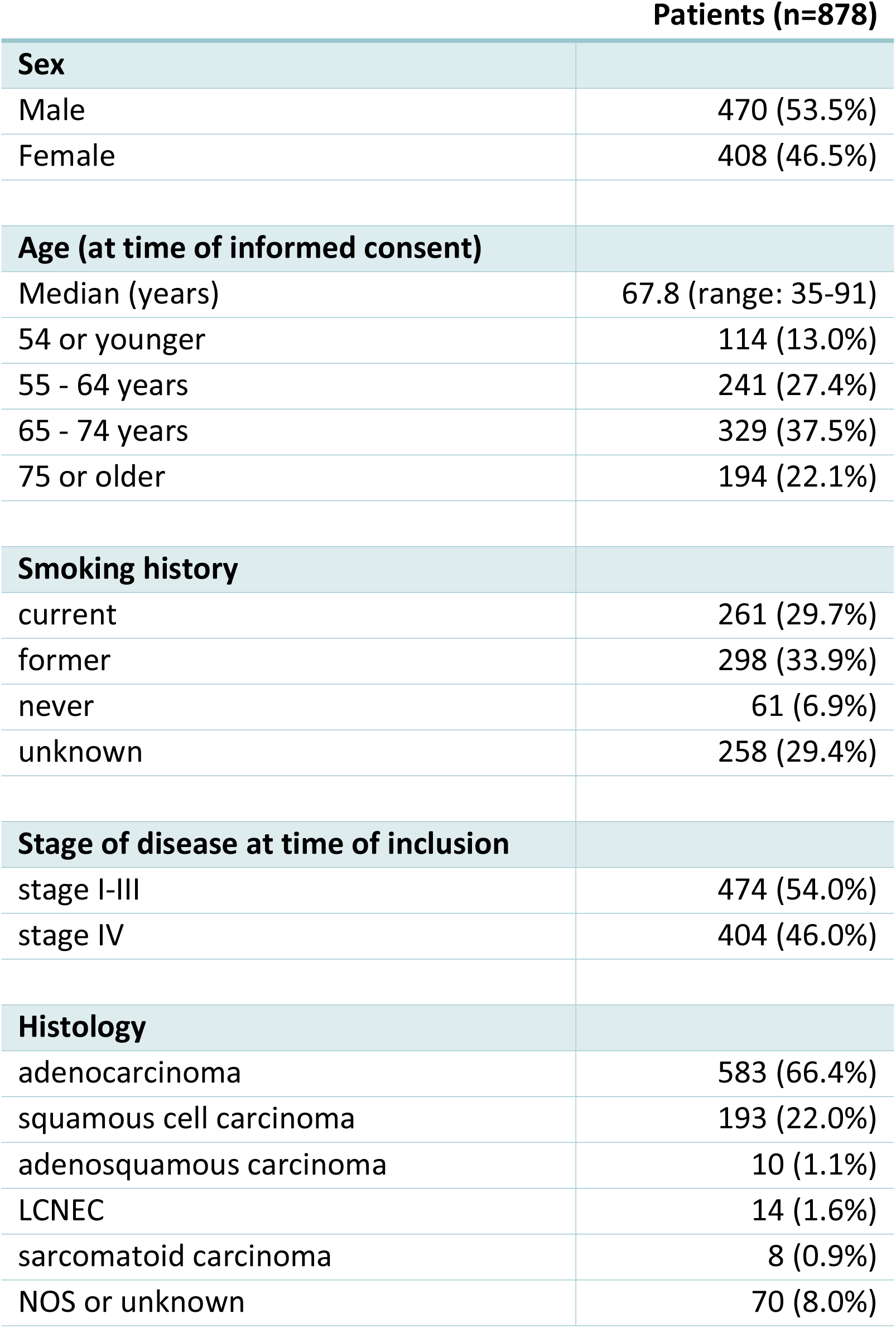
Patient characteristics. Data are n (%) unless stated otherwise. LCNEC = large cell neuro-endocrine carcinoma. NOS = not otherwise specified.

### Tissue-based molecular profiling

#### Quantification of increased testing by study design

In the run-in phase, combined *EGFR* and *ALK* testing was performed in 95 out of 136 patients (69.9%, 95%CI: 61.4%-77.4%) with any stage of NSCLC, significantly more (*p*<0.001) than the estimated 55% in routine clinical care. The overall testing rate further increased to 77.0% (95%CI: 73.8%-80.0%) in the protocolised study phase (*p*=0.049). *EGFR* or *ALK* alterations were detected in 16 patients in the run-in phase (11.8%, 95%CI: 6.9%-18.4%) and in 76 patients in the protocolised phase (10.3%, 95%CI: 8.2%-12.7%. Both detection rates were higher than the postulated 5.8% threshold for clinical relevance (*p*=0.0027 for the run-in phase, *p*<0.001 for the protocolised phase). No statistically significant difference in *EGFR* and *ALK* alteration detection rate was observed (*p*=0.648).

#### *EGFR* and *ALK* profiling rates in subgroups

In all subgroups (stage I-III vs stage IV, and squamous vs non-squamous), the testing rate for *EGFR* and *ALK* increased. In stage I-III the overall testing rate was 55.0% in the run-in phase and 68.6% in the protocol phase. In stage IV the overall testing rate increased from 81.6% to 87.5% (*p*=0.124). Testing rate in any stage non-squamous cell carcinoma was 77.9% in the run-in phase and 82.4% in the protocol phase, compared to 43.8% (run-in) and 57.5% (protocol) in any stage squamous cell carcinoma. The highest testing rate was observed in stage IV non-squamous cell carcinoma in the protocol phase: 91.2%. Detailed overviews of testing per subcategory are shown in figure 2 and supplementary table S1.

**Figure 2.**
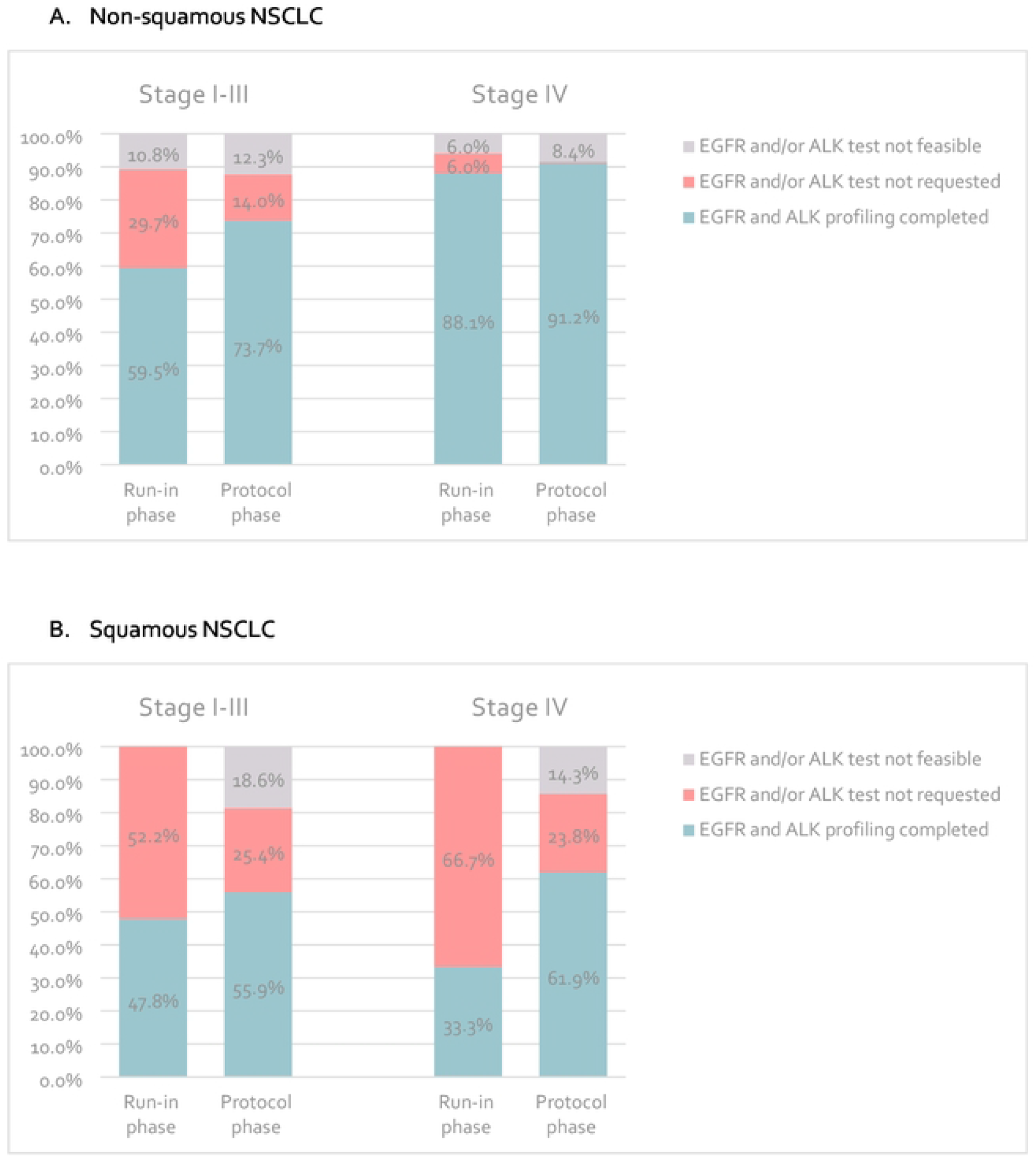
Rates of successful *EGFR* and *ALK* testing. The proportion of patients in whom mutational statusof both *EGFR* and *ALK* were tested in: (A) non-squamous cellcarcinoma. Stage I-Ill: run-in phase n=37, protocol phase n=293. Stage IV: run-in phase n=67, protocol phase n=285. **(B)** squamous cell carcinoma. Stage I-Ill: run-in phase n=23, protocol phase n=118. Stage IV: run-in phase n=g, protocol phase n=42. All absolute numbers and percentages are shownin table S1.

The percentages of patients for whom no *EGFR* and/or *ALK* test was requested decreased from 38.3% to 17.3% in stage I-III (any histology), and from 13.2% to 3.4% in stage IV (any histology), demonstrating the direct effect of protocolising molecular profiling. In any stage squamous NSCLC tumour tissue was insufficient in 17.5% of patients in the protocol phase, compared to 10.4% of patients with non-squamous NSCLC (*p*=0.0186). Reasons for not testing *EGFR* and/or *ALK* are shown in tables 2 and S1.

**Table 2.** Reasons for missing *EGFR* and/or *ALK* tests. The number and proportion of patients in whom mutational status of *EGFR* and *ALK* could not be tested is shown in these tables. Reasons for Numbers indicate n (%). Overall, *EGFR/ALK* profiling was significantly more often unfeasible in squamous cell carcinoma (17.5%, or 28 of 160 patients) than in non-squamous cell carcinoma (10.4%, or 60 out of 578 patients). Based on protocol phase, Fisher’s exact test: *p*=0.0186.

| Non-squamous NSCLC | Stage I-III |  | Stage IV |  |
| --- | --- | --- | --- | --- |
|  | Run-in | Protocol | Run-in | Protocol |
| Patients not tested for *EGFR* and *ALK* | 15 (40.5%) | 77 (26.3%) | 8 (11.9%) | 25 (8.8%) |
| *EGFR* and *ALK* testing not feasible | 4 (10.8%) | 36 (12.3%) | 4 (6.0%) | 24 (8.4%) |
| Insufficient tumour tissue | 3 (8.1%) | 24 (8.2%) | 4 (6.0%) | 15 (5.3%) |
| No tumour tissue obtained | 1 (2.7%) | 8 (2.7%) | 0 | 6 (2.1%) |
| No tumour tissue available at LEMA centre | 0 | 4 (1.4%) | 0 | 0 |
| Patient died before molecular analysis | 0 | 0 | 0 | 3 (1.1%) |
| Squamous NSCLC | Stage I-III |  | Stage IV |  |
|  | Run-in | Protocol | Run-in | Protocol |
| Patients not tested for *EGFR* and *ALK* | 12 (52.2%) | 52 (44.1%) | 6 (66.7%) | 16 (38.1%) |
| *EGFR* and *ALK* testing not feasible | 0 | 22 (18.6%) | 0 | 6 (14.3%) |
| Insufficient tumour tissue | 0 | 12 (10.2%) | 0 | 5 (11.9%) |
| No tumour tissue obtained | 0 | 3 (2.5%) | 0 | 0 |
| No tumour tissue available at LEMA centre | 0 | 6 (5.1%) | 0 | 1 (2.4%) |
| Patient died before molecular analysis | 0 | 1 (0.8%) | 0 | 0 |

#### Exploratory analysis of molecular profiling

Comprehensive molecular profiling (CMP) was defined as testing for oncogenic driver alterations in known oncogenes *EGFR, KRAS, BRAF, ERBB2, ALK, ROS1, RET* and *MET*. CMP data from 3 patients could not be obtained. CMP was successful in 508 out of 739 patients (68.7%) in the protocolised study phase, regardless of histology and stage.

In patients with metastatic NSCLC, CMP was completed in 68.4% during the run-in phase and increased to 79.2% in the protocol phase. (figure 3) Insufficient tumour tissue (quantity or quality), failed biomarker analyses (i.e. not interpretable FISH results), or early death of patients were reasons for incomplete molecular profiling in 9.2% of stage IV patients during the run-in and in 12.2% of patients under study protocol. Incomplete profiling due to missing requests was reduced from 22.4% to 8.6% under the study protocol in stage IV NSCLC. In addition, CMP in stage I-III increased from 51.7% in the run-in phase to 60.4% under the study protocol. (supplementary table S2)

**Figure 3.**
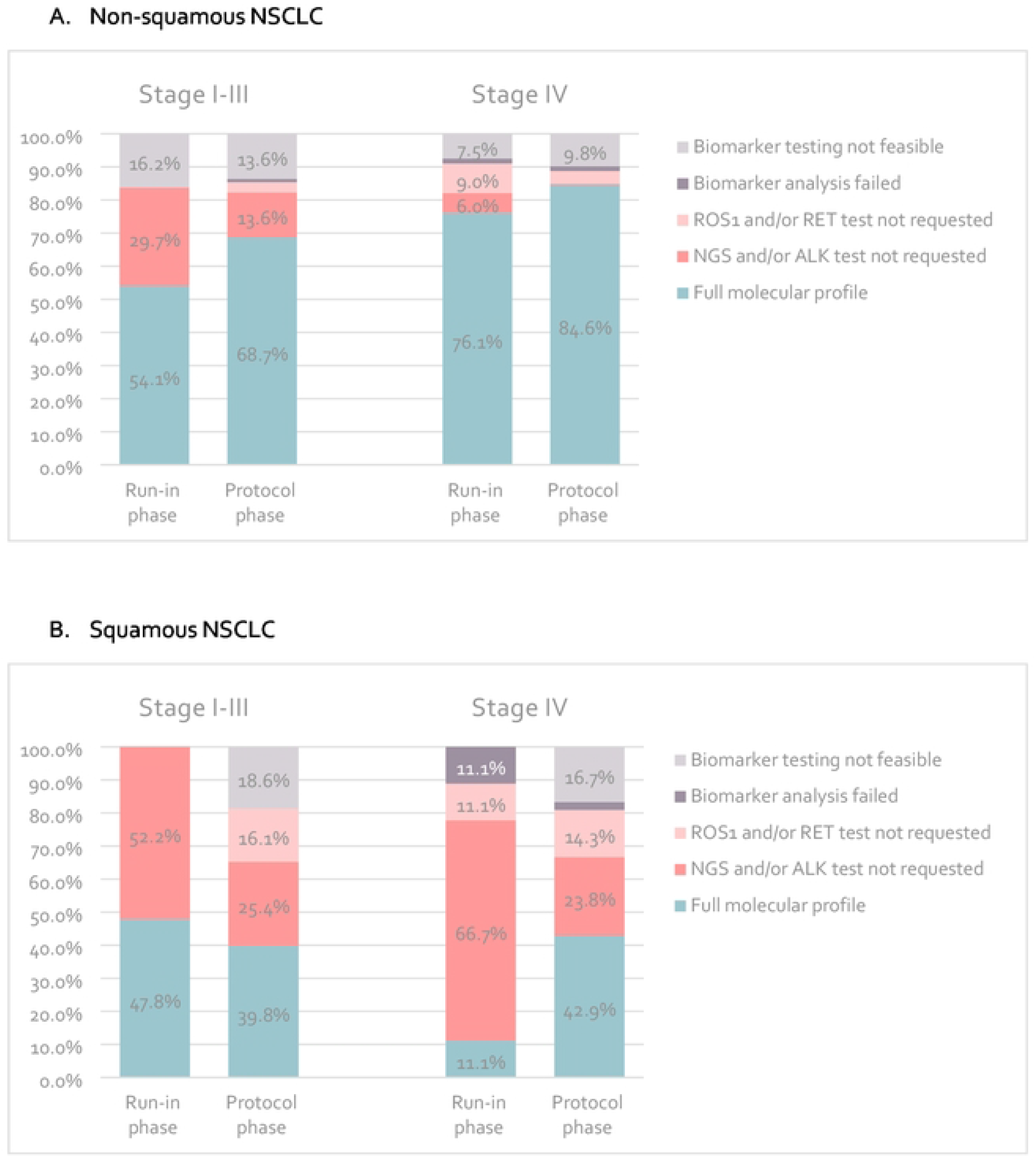
Comprehensive molecular profiling rate. The proportion of patients in whom comprehensive molecular profiling was feasible in: (A) non-squamous cell carcinoma. Stage I-Ill: run in phase n=37, protocol phase n=294. Stage IV: run-in phase n=67, protocol phase n=286. (B) squamous cellcarcinoma. Stage I-Ill: run-in phase n=23, protocol phase n=119. StageIV: run-in phase n=9, protocol phase n=42. All absolute numbers and percentages areshown in table S2.

The extensive molecular profiling of tumours provided insight into the prevalence of both oncogenic driver mutations and PD-L1 expression in various subgroups of NSCLC patients. High levels of PD-L1 expression (>50%) were observed more often in stage IV (approximately 40% of patients) compared to approximately 25% of patients with stage I-III. Targetable driver mutations were seen in all stages and all types of NSCLC. The total prevalence of *EGFR* mutations was 11.3% (14.9% in adenocarcinoma). Total prevalence of *ALK* fusions was 2.5% (3.3% in adenocarcinoma). Supplementary tables S5 and S6 show the prevalence of drivers and the expression of PD-L1 in various subgroups of our cohort.

### Plasma-based molecular profiling

Pre-treatment blood samples were obtained from 821 patients. Thirteen samples were lost in transport or laboratory procedures. Therefore, feasibility of ddPCR for the analysis of *EGFR* or *KRAS* mutations in ctDNA was 98.4%. *EGFR* exon 19 deletions (n=16), *EGFR* L858R mutations (n=16) or *KRAS* codon G12 or G13 mutations (n=100) were detected in 132 patients. Detailed results per patient characteristic are shown in supplementary table S3.

Performance of ddPCR was compared to tissue-based profiling as the gold standard. Sensitivity increased with higher stages of NSCLC and was highest for *EGFR* L858R in stage IV (86.7%). Specificity was consistently above 98% for all subgroups. Test metrics for all assessed alterations of interest in all stages are shown in supplementary table S4. Numbers of detected mutations in tissue, blood or both are shown in figure 4.

**Figure 4.**
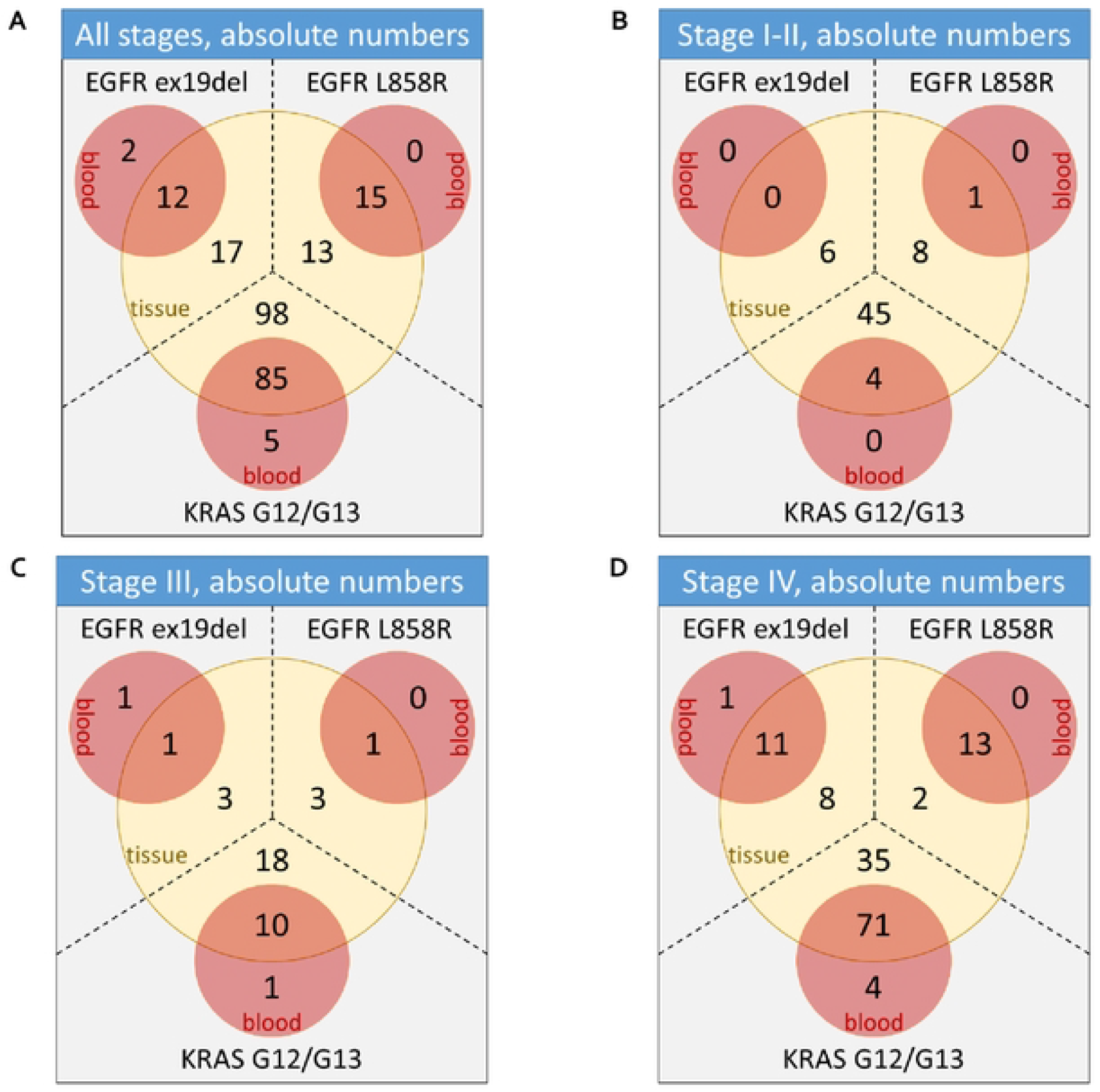
Driver mutations detected by tissue-based analysis, ctDNA analysis, or both. Number indicate the absolute number of samples in which a mutation of interest was detected. (A) mutationsdetected in patients with any stage of NSCLC. (B) mutations detected in patients with stage I-Ill.(C) mutations detected in stage Ill. (D) mutations detected in stage IV.

In patients without tissue profiling *EGFR* ex19del, *EGFR* L858R and *KRAS* G12/G13 mutations were detected by ddPCR in two, one and ten patients, respectively, corresponding to 7.1% of all patients without NGS of tumour tissue. In seven patients with completed tissue-NGS additional driver mutations were found by ctDNA analysis: two *EGFR* exon 19 deletions and five *KRAS* mutations.

## Discussion

This study showed a significant increase in the rate of molecular profiling by protocolising diagnostic routines. Even in metastatic non-squamous NSCLC, where testing is strongly recommended by guidelines, the rate of profiling can be improved. In addition, we have demonstrated that plasma-based profiling is feasible for almost any patient. This method of testing showed reliable test results in stage IV, but sensitivity decreased sharply in earlier stages of NSCLC.

Previously published studies(11,12,18–22) have demonstrated molecular profiling is not performed in a substantial proportion of NSCLC patients. The summarized results in supplementary table S7 show a large variation in the rate of molecular analysis of NSCLC (30.9%-81.8%), although the testing rate increased in recent years, which is also supported by our data. At the start of the LEMA trial, we proposed at least 55% of patients with any stage of NSCLC should be tested for oncogenic alterations in *EGFR* and *ALK*. The testing rate of *EGFR* and *ALK* in the run-in phase of our study was already significantly higher (69.9%, *p*<0.001) than initially expected. This may be explained by increased awareness of local investigators who were already familiarised with the LEMA protocol. The additional increase in testing rate (77.0%, *p*=0.049) demonstrates that further improvement is feasible in real-world clinical practice.

Currently, there is no indication for targeted therapy in early-stage NSCLC. Based on our results no assumption can be made on the clinical benefit following upfront molecular profiling in early-stage NSCLC. However, recent clinical trials suggest that targeted therapy may lead to clinical benefit in early-stage NSCLC as well.(5–7) In addition, molecular profiling data of early stage NSCLC may be used to guide treatment plans in case of disease progression. Therefore, upfront molecular profiling could lead to an optimal personalised choice of treatment for more patients.

We compared our results to the study by Kuijpers(11) et al on data from the Dutch pathology database (PALGA), although this study only included metastatic non-squamous NSCLC. The combined testing rate of *EGFR* and *ALK* in the PALGA study was 61.4% (2013) and 74.7% (2015). The preliminary data from 2017 indicate a further increase in testing. *EGFR* and *ALK* testing rate in our cohort for patients with metastatic non-squamous NSCLC was 91.2%. The large improvement is most likely explained by the central guidance of local laboratories and oncologists. In almost all patients without molecular profiling in the LEMA cohort, this was due to a lack of suitable tissue.

Furthermore, with an increasing number of targetable mutations that are relevant in treatment decision making in NSCLC, extensive testing will be needed. This increases the need for more tumour tissue and decreases the feasibility of CMP. The directive upfront approach in the LEMA study increased the rate of CMP in stage IV NSCLC from 68.4% in the run-in phase to 79.2% in the protocol phase. However, tissue-based CMP will remain unfeasible in a minority of patients due to insufficient tumour tissue, or in patients whose clinical condition does not allow for an invasive procedure.

We have demonstrated that treatable targets are present in various subtypes of NSCLC, but it is evident that molecular profiling of squamous cell carcinoma is less likely to result in the detection of targetable alterations. Furthermore, profiling of early-stage NSCLC is not yet recommended by guidelines and may also be regarded as overdiagnosis. However, adjuvant treatment opportunities are emerging and besides, those patients with early stage disease who finally progress might benefit from upfront molecular profiling, thereby preventing further diagnostic delays. Additionally, the higher rate of molecular profiling comes with additional costs of the diagnostic work-up. The financial burden of extensive testing must be weighed against the benefits of more patients receiving their optimal treatment. The cost-effectiveness of the approach of the LEMA study is still to be determined by health technology assessment.

In addition, this study demonstrates that ctDNA ddPCR analysis on a limited volume of plasma is feasible in almost all patients and detects oncodriver mutations in patients without tissue profiling, but also additional mutations in patients with already completed tissue profiling. Sensitivity and specificity compared to tissue analysis were in line with previously published results and were promising for clinical use in stage IV NSCLC.(23–25) The utility in earlier stages of disease is questionable, given the substantially lower sensitivity. This may be explained by lower quantities of ctDNA in the blood of patients with a lower tumour burden. The performance of ctDNA analysis by ddPCR in our cohort showed additional value of this test next to tissue analysis in the diagnostic work-up of metastatic NSCLC.

## Conclusion

This study demonstrates comprehensive molecular profiling can be optimised by a protocolised approach. However, tissue-based profiling will remain unfeasible for a minority of patients. A combination of tissue- and plasma-based analyses is a promising approach to enable molecular profiling in more patients, given the favourable test characteristics of ctDNA analysis in the metastatic setting.

## Data Availability

Data are available from the Netherlands Cancer Institute Institutional Data Access (contact via) for researchers who meet the criteria for access to confidential data.

## Acknowledgements

Special thanks to M. Muller^1^, D. Vessies^1^, E. Platte^1^, M. Lucas^1^, T. Korse^1^, R. van der Wiel^1^, L. Geurts^2^, A. Olijve^3^, A. Van Groensteyn^4^, J. Ippel^5^, A. Moons-Pasic^6^, N. Dijkstra^6^, M. Manschot^7^, A.J. van der Wekken^8^ and H.J.M. Groen^8^ for their efforts in making this project possible and for their valuable contributions to this manuscript.

^1^ Netherlands Cancer Institute, Amsterdam

^2^ Radboud University Medical Centre, Nijmegen

^3^ Noordwest Ziekenhuis Groep, Alkmaar

^4^ University Medical Centre Utrecht

^5^ Haaglanden Medical Centre, The Hague

^6^ Onze Lieve Vrouwe Gasthuis, Amsterdam

^7^ Tergooi Ziekenhuizen, Hilversum

^8^ University Medical Centre Groningen

All centres are located in the Netherlands

